# Antibody response in individuals affected with Sars-Cov-2 infection: temporal trends and qualitative and quantitative differences in symptomatic and asymptomatic subjects. A Cross Sectional Analysis. Ab-Covid Study

**DOI:** 10.1101/2021.08.09.21261450

**Authors:** Iosief Abraha, Antonella Germani, Erica Pasquarelli, Sofia Pascolini, Rossana Antonietti, Sandro Argenti, Alessandra Fioravanti, Elisa Martini, Luana Aristei, Paola Mancinelli, Maria Letizia Ottaviani, Martina Roselli, Milena Barzacca, Erika Belardinelli, Marta Micheli

**Affiliations:** Servizio Immunotrasfusionale, Azienda USL Umbria 2, Foligno

**Author notes:** Correspondence to: Dr. Iosief Abraha.

## Abstract

**Objectives:** To describe clinical characteristics and treatment used in subjects who had Sars-Cov-2 infection during the first pandemic and to assess the correlation between serological titers and clinical characteristics; to evaluate the persistence of antibody titer.

**Design:** Cross-sectional study; 12 months follow-up.

**Setting:** Residents in Azienda USL Umbria 2.

**Participants:** Consecutive subjects aged 15 to 75 who were discharged with the diagnosis of Sars-Cov-2 from the hospitals of the AUSL Umbria 2, or resulted positive to a PCR test for Sars-Cov-2 infection with or without symptoms. SARS-CoV-2 serologic testing for antibodies targeting the Nucleocapside and Spike proteins were determined.

**Results:** Of 184 eligible subjects, 149 were available for evaluation: 17 were classified as Oligo/asymptomatic, 107 as Symptomatic, 25 as Hospital admitted. While fever resulted common to all the groups, headache or musculoskeletal pain was common to symptomatic participants whereas cough and dyspnea was present in all the hospital admitted. Participants with significant signs and symptoms were more likely to use antibiotics, hydroxychloroquine, heparin and steroids. Compared to Oligo/asymptomatic participants, Symptomatic and Hospital admitted participants had higher levels of anti-S titers at every follow-up (median titer at 12 month follow-up: 29 vs 94 vs 116 respectively; P < 0.001). At 12 months follow-up, anti-S titers persisted above the threshold for at least 12 months in all Hospital admitted participants, in 90% of the Symptomatic participants and 83% in the oligo/asymptomatic participants; in 30% of participants the titer raised significantly probably due to reinfection. Anti-N antibody titer tended to decrease over time and in 62% of the entire cohort resulted negative. None of the participants reported clinical reinfection with Sars-Cov-2 virus.

**Conclusion:** Anti-S and anti-N antibody titers correlates well with disease severity. Anti-S antibodies persist for at least one year and most probably provide protection from reinfection.

**Strengths and limitations of this study**

- The key strength of this study is the evaluation of anti-Sars-Cov-2 serology using two types of serological assays and the follow-up that endured for at least 12 months
- In addition to serological evaluation participants were also followed-up clinically
- The study does not have a baseline serologic testing since it was conceived in late April when most of the participants were discharged from hospital or had their symptoms resolved
- The study lacks clinical and serological information regarding those who died during the pandemic event, hence we are unable to conclude whether quantitative serologic testing could predict survival

## Background

The novel acute respiratory syndrome coronavirus 2 (SARS-CoV-2) virus has caused a pandemic infection known as COVID-19. The disease is associated with severe morbidity and mortality, currently threatening global health as well as economy. The disease presents important challenges in different settings including prevention, treatment as well as diagnostic and prognostic significance based on immune response.

The diagnosis of COVID-19 disease depends much on clinical or epidemiological context though it is mainly based on the molecular testing of symptomatic subjects. However, false-negative PCR results are not infrequent and a significant proportion of infected people might remain asymptomatic [1-4]. Unlike the nasopharyngeal RT-PCR tests, the antibody tests allow for better collection of epidemiological data, determination of the immune status of asymptomatic individuals, and screening of previous exposure [5]. Hence, Covid-19 serologic tests, despite their limitation and somewhat challenging performance characteristics, can be an appropriate tool to better diagnose recent or past infection [4]. Use of antibody testing in the context of SARS-CoV-2 infection is being encouraged also to assess the presence of immunity for SARS-CoV-2 infection, to prevent the spread of the diseases [6] as well as to identify people with secure immunity in order to make them return to work [7]. Additionally, serological diagnosis is becoming an important tool to understand the extent of COVID-19 in the community [8] including to estimate true prevalence of the disease. A cross-sectional study in a random sample of blood donors estimated showed that 4.6% of healthy adults were already positive for Sars-Cov-2 antibodies[9].

Studies showed that within 5 days of Sars-Cov-2 infection, the IgM antibodies increase from 50% to 81%, whereas the IgG antibodies increase from 81% to 100%[10, 11]. World Health Organization guidelines recommend obtaining a blood sample during the first week of illness and then 3 to 4 weeks later to measure SARS-CoV-2 antibodies[11, 12].

Serologic tests have been introduced to detect antigens namely the spike protein (S), the protein nucleocapside (N) and the virus membrane[6]. N and S proteins were found to be the major immunogenic proteins [13]. As in the MERS-CoV infection, antibodies against proteins S, 3a, N, and 9b were detected in the sera from convalescent-phase SARS patients [13]. Though anti-S and anti-N were dominant and could persist in the sera of SARS patients until week 30, only anti-S3 demonstrated significant neutralizing activity [13].

Current methods available for serologic testing include rapid diagnostic tests (RDT), enzyme-linked immunosorbent assays (ELISA), neutralization assays, and chemiluminescent immunoassays (CLIA) [6]. ELISA and CLIA are considered suitable for first line screening because of the large throughput, short processing time, and simple operating procedure [11].

A Cochrane review evaluated the diagnostic accuracy of antibody tests to determine whether a person presenting in the community or in primary or secondary care has SARS-CoV-2 infection, or has previously had SARS-CoV-2 infection [14]. The reference standards for comparing these antibody tests were RT-PCR tests and clinical diagnosis based. After excluding several studies due to relevant bias, the authors analyzed 19 studies and concluded antibody tests are likely to have a useful role for detecting previous SARS-CoV-2 infection if used 15 (sensitivity 91%; 95% CI 87 to 94) or more days after the onset of symptoms [14].

In addition to antibody profile, longitudinal persistence of immunity in convalescent Covid-19 subjects has been another issue of debate for months after the first pandemic. An observational study published during that pandemic [15] found in 23 patients a positive correlation between enzyme immunoassay antibodies and neutralizing antibody titer but concluded that further investigation is needed on the role of anti-COVID antibodies in immunopathology and / or antiviral treatment [15].

We performed a longitudinal cohort study in Umbria of subjects with a confirmed diagnosis of Sars-Cov-2 between February and April 2020 with a follow-up of at least 12 months. Levels of IgG antibodies against SARS-CoV-2 Nucleocapside (N), and neutralizing antibodies were determined.

The objectives of our study was to describe clinical characteristics and treatment used in subjects who had Sars-Cov-2 infection during the first pandemic; to assess the correlation between serological titers and clinical characteristics; to evaluate the persistence and trend of anti-Sars-Cov-2 titers over a follow-up of 12.

## Methods

### Study design and target population

The present study was a cross-sectional in design. Our cohort of interest was characterized of consecutive subjects aged 15 to 75 who from February, 2020, to April 2021, (a) were discharged with the diagnosis of Sars-Cov-2 from the hospitals of the AUSL Umbria 2, or (b) resulted positive to a PCR test for Sars-Cov-2 infection. These subjects were invited to undertake a serologic SARS-CoV-2 testing for antibodies targeting the Nucleocapside (N) protein and S proteins of SARS-CoV-2. All the cohort was clinically and serologically followed-up longitudinally. After enrollment, serology testing were performed every 3-4 months for every participants until end of follow-up. Clinical signs and symptoms as well as specific COVID-19 treatments were recorded at baseline and during follow-up.

The planning conduct and reporting was performed in accordance with the Declaration of Helsinki, as revised in 2013. This study was approved by the Regional Research Ethics Committee (Comitato Etico Regionale – Umbria, CER 3695/20), and written informed consent was obtained.

### Laboratory methods

Serum samples were analyzed using two commercial serologic assays: Abbott SARS-CoV-2 IgG, DiaSorin Liaison SARS-CoV-2 S1/S2 IgG.

The qualitative detection of anti-N IgG was performed using a chemiluminescent microparticle immunoassay (Abbott ARCHITECT SARS-CoV-2 IgG). A signal/cut-off ratio of ≥1.4 was interpreted as reactive according to the manufacturer’s instructions [16]. Studies report that clinical sensitivity is time-dependent and after day-14 it ranges between 84.2–100% whereas specificity results 99.6%-100%[17, 18]. Prior to analyses of patient samples, calibration was performed and negative quality control signal/cut-off ratio ≤ 0.78 and positive quality control signal/cut-off ratio 1.65–8.40 were achieved.

The quantitative detection of anti-S IgG was evaluated using a standardized automated chemiluminescent assay (DiaSorin S.p.A., Saluggia, Italy). A detection of ≥ 12 AU/ml was interpreted as positive according to the manufacturer’s instructions [19]. The test’s sensitivity is time-dependent, that is 25% in the first 5 days after RT-PCR-confirmed diagnosis, 90% from day 5 to day 15, and 97% from day-15 forward.

### Statistical analysis

Demographic characteristics of the study participants was described by calculating the frequencies and percentages for categorical variables, and medians and inter-quartile intervals for continuous variables. The analysis of the normal distribution of the sample was evaluated using the Kolmogorov - Smirnov test using STATA software, with the significance level at P <0.05.

To compare antibody titers among the study groups analysis of variance (ANOVA) was planned for comparison of means when parametric criteria were reached; for non-parametric distributions, we used the U-Mann–Whitney and Kruskal–Wallis tests. We used the regression analysis to adjust for statistical significance for multiple comparisons and to account for potential confounding factors (e.g., age,sex).

Differences between proportions were evaluated by χ^2^ tests. We calculated 95% CIs, and P values <0.05 were considered statistically significant throughout the analysis. No imputation was performed for missing data.

Patients or the public were not involved in the design, or conduct, or reporting, or dissemination plans of our research

## Results

184 potentially eligible subjects were identified. After excluding 35 subjects with reasons 149 met the inclusion criteria and signed the informed consent. Of this cohort 21 were not available to perform the serologic test at the 2^nd^ follow-up but 19 of these returned for the last follow-up.

Subsequently, 14 subjects received anti-Covid vaccination and 6 were unavailable for serologic testing and were excluded from analysis. At 12 months follow-up 130 participants were still available for clinical and serologic evaluation. All of the excluded subjects at final follow-up were traceable through telephone contact and were possible to obtain the their health status. **Figure 1** shows the study screen process.

**Figure 1.**
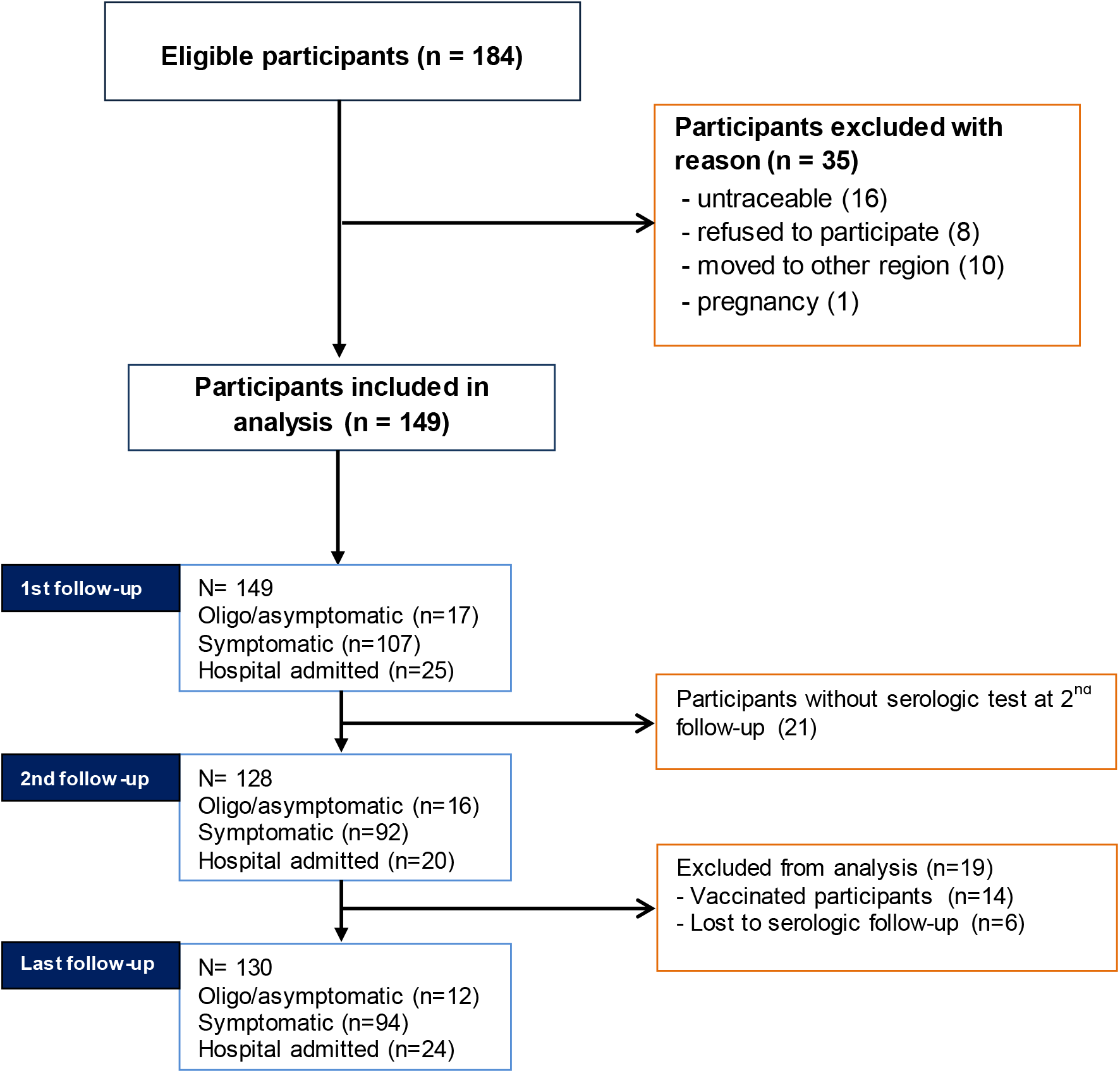
Study screening process

Of the initial cohort, 17 (11%) were oligo/asymptomatic, 107 (72%) were symptomatic participants (without hospital admission), 25 (17%) were participants who were admitted to hospital. The mean age was 49 years (median 54). While 52% of the cohort were female, men tended to have more severe symptoms reaching 80% of the Hospital admitted participants (**Table 1**).

**Table 1.** Basic characteristic of the cohort classified by symptom severity.

|  | Oligo/asymptomatic participants | Symptomatic participants without hospital admission | Hospital admitted participants |
| --- | --- | --- | --- |
| N (%) | 17 (11) | 107 (71) | 25 (16) |
| Male (%) | 11 (64) | 40 (37) | 20 (80) |
| Age (median; p25, p75) | 42 (33 – 57) | 53 (39 – 59) | 56 (54-64) |
| **Clinical signs and symptoms** |  |  |  |
| Fever | 7 (44) | 94 (83) | 23 (92) |
| Headache/musculoskeletal pain | 3 (19) | 58 (51) | 11 (44) |
| Ageusia/anosmia | 7 (43) | 59 (52) | 3 (12) |
| Asthenia | 1 (6) | 59 (52) | 8 (32) |
| Cough | 0 (0) | 43 (38) | 23 (92) |
| Dyspnea | 0 (0) | 17 (15) | 25 (100) |
| Pneumonia | 0 (0) | 3 (3) | 25 (100) |
| **Treatment** |  |  |  |
| Antibiotic | 0 (0) | 28 (25) | 25 (100) |
| Hydroxychloroquine | 0 (0) | 12 (10) | 25 (75) |
| Heparin | 0 (0) | 1 (1) | 24 (96) |
| Antiviral | 0 (0) | 0 (0) | 8 (32) |
| Monoclonal antibody | 0 (0) | 0 (0) | 4 (16) |
| Steroids | 0 (0) | 14 (13) | 5 (20) |
| NSAIDS | 0 (0) | 7 (6) | 0 (21) |
| Paracetamol | 1 (6) | 20 (18) | 1 (4) |

### Type and duration of symptoms

The most common symptom was fever which resulted common to all the three groups. Headache or musculoskeletal pain was common to symptomatic participants whereas cough and dyspnea was present in all of the admitted participants indicating the severity of the disease. All hospital admitted participants had radiographically documented pneumonia (**Table 1**).

The most persistent symptoms were asthenia (median 30 days) as well as anosmia and/or ageusia (median 30 days). Anosmia/ageusia persisted across the three groups and the median symptoms’ duration increased as severity of symptoms increased (median: 6 days in Oligo/asymptomatic participants, 20 days in Symptomatic participants, 30 days in Hospital admitted participants).

Similarly, median duration for asthenia was 20 days in the Oligo/asymptomatic participants, 30 days in Symptomatic participants, and 25 days in Hospital admitted participants. In 35 patients, anosmia/ageusia lasted for more than 6 months but resolved completely within 10 months. Duration of symptoms across the three groups of participants are depicted in **Figure 2**. Duration of ageusia/anosmia and asthenia resulted higher in females than in males.

**Figure 2.**
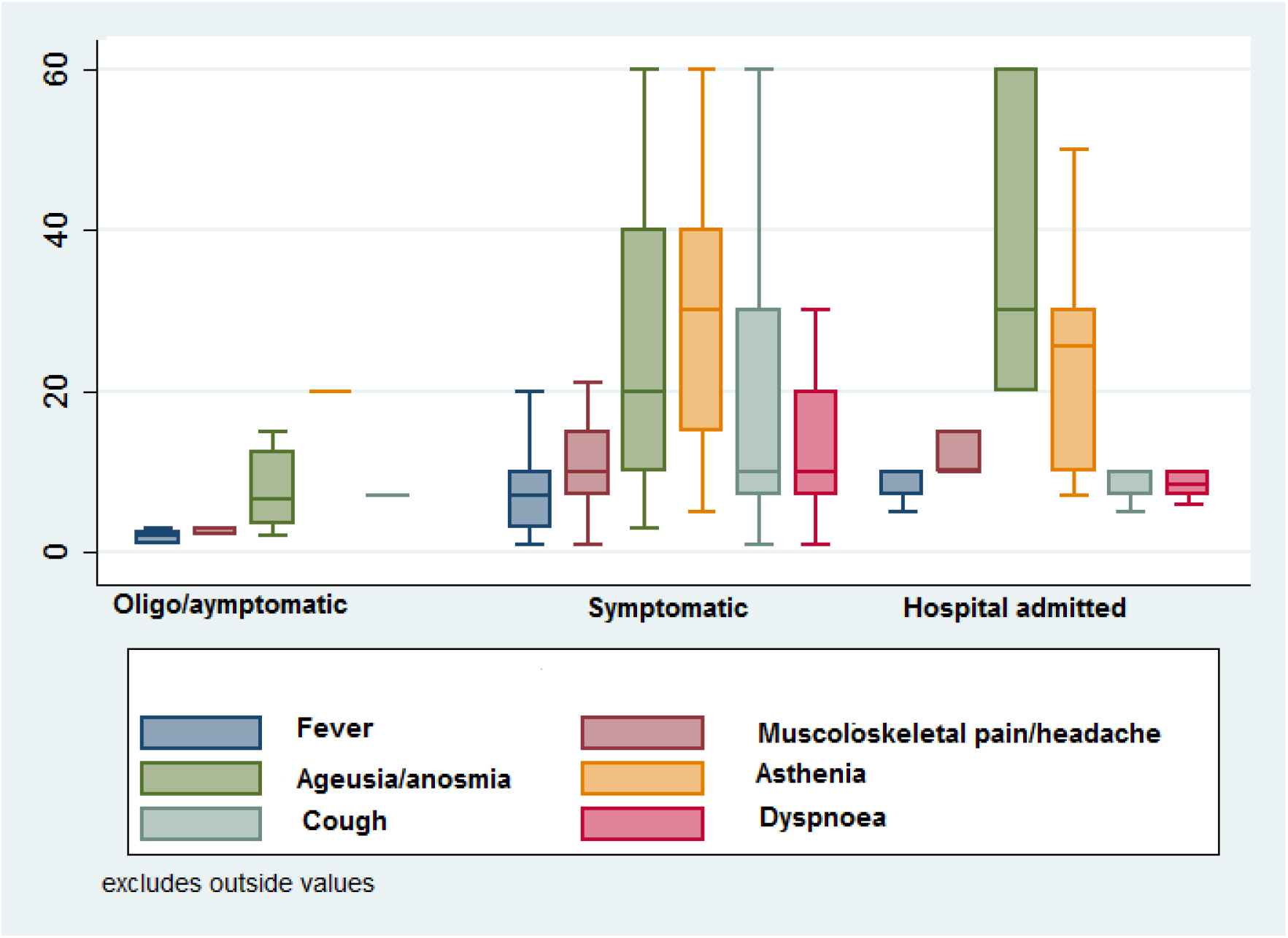
Duration of signs and symptoms across the three groups of participants.

**Figure 3.**
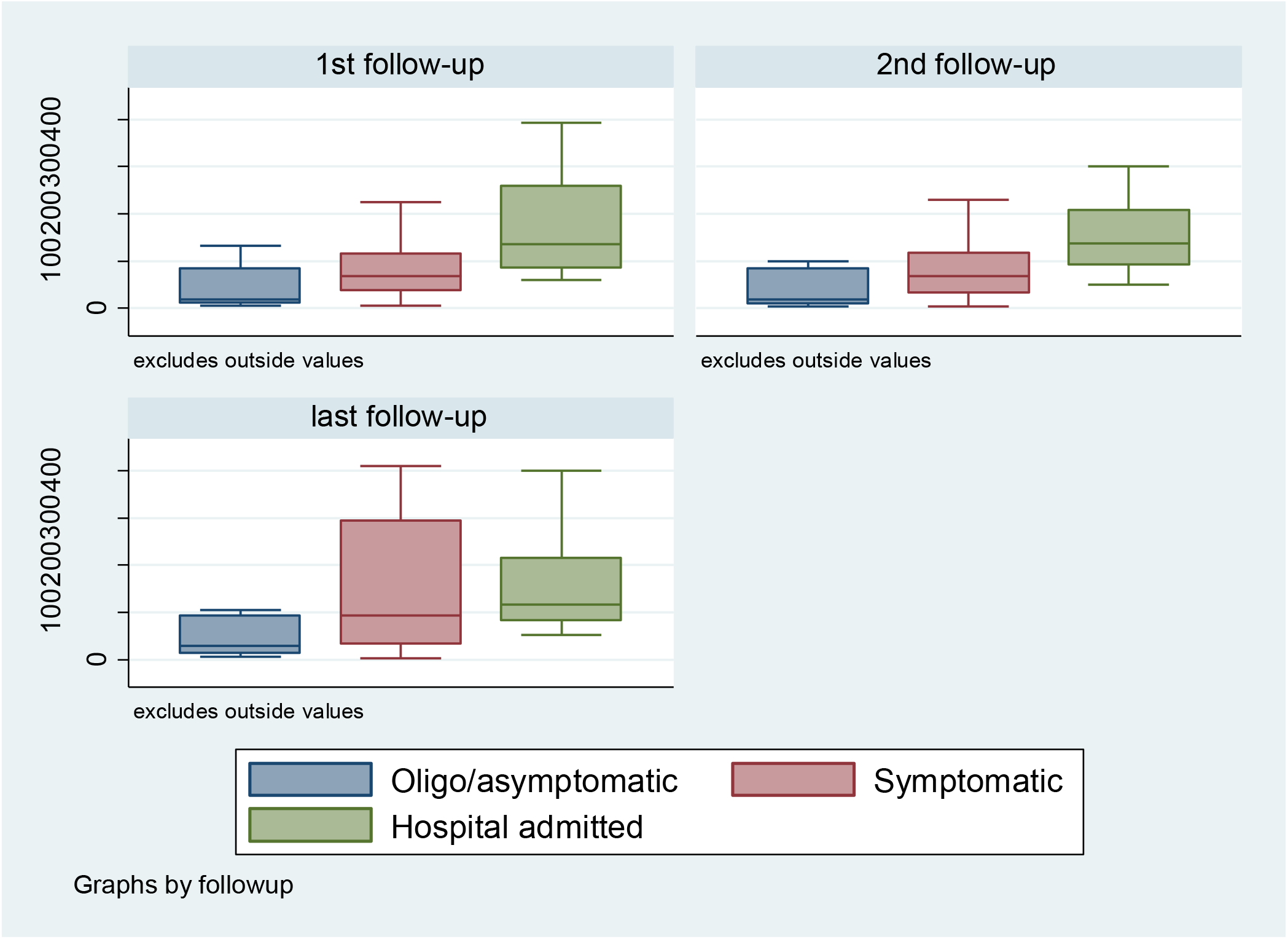
Ab-anti-S titer across the three Groups of participants compared between the three periods of follow-up.

### Treatment used

Most of the Oligo/asymptomatic participants were not treated or reported the use of paracetamol or anti-inflammatory agents. Anti-inflammatory agents were most used in Symptomatic participants. Twenty-five percent of Symptomatic participants and 100% of Hospital admitted participants used antibiotics. Hydroxychloroquine was used by 10% of the Symptomatic participants and by 100% of Hospital admitted participants. Low-dose heparin was almost exclusively used by hospitalized participants. Antivirals and monoclonal antibodies were used in the 32% and 16% of the hospitalized patients, respectively (**Table 1**).

### Serological outline – Anti-Spike

The median value of the antibody anti-S (Diasorin) titer at first visit (that is between June and September 2020) across the whole cohort was 74 U/ml (IQR 92). The median anti-S titer was 74 U/ml, (IQR 105) in males and 71 U/ml (IQR 83) in females.

Anti-S antibody values differed significantly across the three groups of participants. At first time follow-up, median titers in the symptomatic and hospital admitted participants were significantly higher compared to the oligo/asymptomatic participants; similarly, anti-S titer levels were higher in the Hospital admitted participants compared to Symptomatic subjects indicating that the more significant were the clinical signs and symptoms the higher was the anti-S antibody response. At subsequent follow-up the median titer of anti-S antibodies resulted substantially similar with respect to the values observed at first visit and the statistical difference within groups remained constant overtime (**Table 2**).

**Table 2.** Median (interquartile range) Anti-N and Anti-S titers according to clinical classification of participants

|  | Oligo/asymptomatic participants | Symptomatic participants | Hospital admitted participants | Non-parametric test |
| --- | --- | --- | --- | --- |
| **Anti-Spike serology** |  |  |  |  |
| *1<sup>st</sup> follow-up*<br>*N=149* | 18 (12, 85) | 68 (36, 115) | 135 (84, 259) | P < 0.001 |
| *2<sup>nd</sup> follow-up*<br>*N=128* | 16 (10, 85) | 69 (31, 118) | 138 (93, 208) | P < 0.001 |
| *Last follow-up*<br>*N=130* | 29 (16, 92) | 94 (33, 205) | 116 (81, 216) | P < 0.001 |
| **Anti-Nucleocapside serology** |  |  |  |  |
| *1<sup>st</sup> follow-up*<br>*N=149* | 2.31 (0.62, 3.87) | 3.03 (1.26, 4.77) | 4.55 (2.89, 6.02) | P = 0.094 |
| *2<sup>nd</sup> follow-up*<br>*N=128* | 0.8 (0.34, 2.99) | 1.32 (0.54, 2.44) | 2.35 (1.75, 4.22) | P = 0.056 |
| *Last follow-up*<br>*N=130* | 0.34 (0.23, 0.88) | 0.77 (0.33, 1.81) | 1.32 (0.69, 2.97) | P = 0.36 |

In addition, percentage difference between the first and second follow-up and between the first and last follow-up were calculated where antibody titers were considered increased (or decreased) when there was at least a 10% increase (or decrease) in the percentage difference. Between the first and second follow-up there was an increase in the anti-S titer by 35% in each of the symptomatic and hospital admitted participants in contrast to 19% in the oligo/asymptomatic participants. During this period of observation in 25% of the overall cohort the anti-Spike titer remained constant maintaining a range between -10% and +10% (**Table 3**).

**Table 3.** Percentage difference and median of anti-Spike and anti-Nucleocapside titers between first and second follow-up, between first and last follow-up

|  | Oligo/asymptomatic participants | Symptomatic participants | Hospital admitted participants | All |
| --- | --- | --- | --- | --- |
| **Anti-Spike antibody difference between first and second follow-up** |  |  |  |  |
| **Median (IQR)** | -9.5 (21.4) | 0 (46.7) | -10 (57.1) | -2 (50.8) |
| **>10** | 3 (19) | 32 (35) | 7 (35) | 42 (33) |
| **- 10 to +10** | 6 (38) | 23 (25) | 3 (15) | 32 (25) |
| **< -10** | 7 (44) | 37 (40) | 10 (50) | 54 (42) |
| **Anti-Spike antibody difference between first and last follow-up** |  |  |  |  |
| **Median (IQR)** | -19.7 (52.5) | 9 (88.8) | 9.3 (43.7) | 8.1 (78.6) |
| **>10** | 2 (17) | 44 (47) | 12 (50) | 52 (45) |
| **- 10 to +10** | 5 (42) | 19 (20) | 5 (21) | 22 (22) |
| **< -10** | 5 (42) | 31 (33) | 7 (29) | 39 (33) |
| **Anti-Nucleocapside antibody difference between first and second follow-up** |  |  |  |  |
| **Median (IQR)** | -65 (69) | -105 (144) | -87.9 (102) | -100 (123) |
| **>10** | 4 (25) | 6 (7) | 1 (5) | 11 (9) |
| **- 10 to +10** | 1 (6) | 6 (7) | 2 (10) | 9 (7) |
| **< -10** | 11 (69) | 80 (87) | 17 (85) | 108 (84) |
| **Anti-Nucleocapside antibody difference between first and last follow-up** |  |  |  |  |
| **Median (IQR)** | -366 (550) | -217 (336) | -222.7 (317) | -220 (352) |
| **>10** | 0 (0) | 1 (1) | 0 (0) | 1 (1) |
| **- 10 to +10** | 2 (17) | 18 (19) | 4 (17) | 24 (18) |
| **< -10** | 10 (83) | 75 (80) | 20 (83) | 105 (81) |

Interestingly, difference between the first and last follow-up showed an increase in the anti-S titer in around 47% of the symptomatic participants and 50% of the Hospital admitted participants in contrast to 17% of the Oligo/asymptomatic participants. During the whole period of observation 67% the anti-S titer remained stable or increased (**Table 3**).

The decrease of the difference in percentage of the titer between the first and the last follow-up by at least 10% was observed in 33% of the whole cohort. Nonetheless the subjects that showed an anti-S titer below the threshold of 12 U/ml was less than 10% of the available cohort: 9.4% at first follow-up, 8,6% at the second and 9,2% at the third. Most of these events occurred in the oligo/asymptomatic participants whereas none of the hospital admitted subjects had their antibody titer below threshold (**Table 3**).

### Serological outline – Anti-Nucleocapside

The median value of the antibody anti-N titer at first visit across the whole cohort was 3.1 U/ml (IQR 3.63) with non-significant higher values in males (3.75 U/ml, IQR 3.93) than females (2.46 U/ml, IQR 3.33).

Antibody values were higher in the hospital admitted participants compared to the other two groups at every follow-up time but with no statistically significant difference (**Table 2**). In addition, in every group of participants the antibody titer reduced constantly overtime. When anti-N titer was compared within groups, there appears to be a downward trend across the groups during the whole period of follow-up.

Difference in percentage of anti-N titer between the first and second follow-up and between the first and last follow-up showed a substantial decrease in the serologic titer across the three groups of participants (**Table 3**). The percentage of the subjects with serologic titer under threshold (< 1.4 U/ml) increased from 26% to 62%. This increase was higher in the oligo/asymptomatic and symptomatic participants (**Table 4**).

### Clinical follow-up

During serologic follow-up participants underwent a clinical visit. When participants were not available for clinical visit their health status was ascertained through telephone call. Particular attention was provided to those who had their anti-S titer augmented. None of the participants in any of the group had any sign or symptom that could be attributed to a possible Sars-Cov-2 reinfection.

## Discussion

We enrolled a substantial number of subjects to whom a diagnosis of Sars-Cov-2 was made during the first pandemic episode within the area of Local Health Unit 2 of Umbria where the main hospitals to which participants had access were Foligno, Spoleto and Orvieto. Participants were invited to sign a consent and to undergo a serologic test, together with a clinical visit every three to four months. Participants were classified according to clinical severity to asymptomatic or paucisymptomatic group, symptomatic subjects with no history of hospital admission and symptomatic subjects with history of hospital admission. Paucisymptomatic subjects were defined according to symptoms that lasted for less than 3 days or when only a non-acute symptom (ageusia-anosmia or asthenia) persisted for less than 15 days. Participants that were admitted to hospital had more severe symptoms that include persistent cough and dyspnea and had radiologically ground glass interstitial pneumonia.

Our study showed also that anti-S antibody response was significantly higher in patients who had noteworthy symptoms. In particular, subjects that were admitted to hospital or had documented pneumonia showed significant levels of antibody titer with a median that was higher than 100 U/ml compared to participants who belong to the other two groups. These results are in agreement with several studies published in medical literature. Compared to hospitalized patients with severe illness, nonhospitalized patients with mild disease typically have lower levels of antibodies than hospitalized patients with severe illness[20-22]. The highest antibody titers observed in severely ill participants might explained in that severe disease is associated with uncontrolled inflammation and significant viral replication stimulates excessive production of antibodies. A study that evaluated antibody responses in 113 patients with Sars-Cov-2 found that the most severely affected patients, in addition of exhibiting high anti-spike antibody levels, showed also the highest levels of inflammatory markers and pro-inflammatory cytokine signatures[23].

Differences in antibody response between individuals may be determined also in part by differences in antigen exposure, age and gender. In a study that evaluated humoral immune response in 126 potential convalescent plasma donors, the authors found that male sex, older age, and hospitalization for COVID-19 were associated with increased antibody responses across the serological assays[24]. In our study most of the subjects that were hospitalized were males and they had a median age that was higher than in Symptomatic participants or Oligo/asymptomatic participants. However, we are unsure whether other factors such as time of diagnosis, sampling efficiency using swab samples, and early treatment – such as steroids[25] – might have influenced the intensity of antibody responses[23].

We found also that in most of the participants that exhibited anti-S at the initial screening the antibody titers persisted for 12 months with decrease of the anti-S below threshold in less than 10% that were mostly asymptomatic participants. Despite initial reports that persistence of antibody against SARS-Cov-2 was limited to a few months[26] and that recovered individuals are prone to reinfection [27], subsequent studies reported that antibodies against SARS-CoV-2 persist over time. In a population-based study performed in Iceland, 15% of the country’s population (around 30,000 individuals) was tested for infection with SARS-CoV-2 by quantitative PCR and antibody testing. Importantly, anti-N and anti-S antibodies remained stable over the 4 months after diagnosis**[28]**. In another cohort of more than 30,000 infected individuals with mild to moderate COVID-19 symptoms, Wajnberg *et al*. assessed the robustness and longevity of the anti–SARS-CoV-2 antibody response. After demonstrating that anti-spike binding titers significantly correlate with neutralization of authentic SARS-CoV-2, the authors showed that antibody titers remain relatively stable for at least a period of about 5 months[29]. More recently, Zhenyu He and colleagues [30] report their cross-sectional study of serological responses of more than 9500 individuals from 3600 households in Wuhan, the first epicenter of the Covid-19 disease. In this study neutralizing antibodies developed in approximately 40% of antibody-positive individuals. In this subgroup the proportion of participants who were positive for IgG and neutralizing antibodies, and the titers of neutralizing antibodies, did not significantly decrease during the 9 months of observation. Similarly, Favresse *et al*, found stable antibody titers over a period of 10 months with the highest positivity rates in patients with clinically significant past SARS-CoV-2 infection[31].

These results are in agreement with ours that showed anti-Spike antibodies persisted for at least one year in the subjects that showed infection between February and March 2020 and resulted serologically positive. In addition to this important finding, our results showed also that 32% of participants serologically positive at the initial visit, reported a significant increase in the antibody titers during follow-up and this increase occurred during subsequent pandemic infections. Importantly, none of these participants reported reinfection indicating that these antibodies have a protective effect on Sars-Cov-2 infection. In general, participants that exhibited increase in antibody titers were working on public (e.g., pharmacists, nurses, medical doctors, bus drivers etc.) and were more likely to be re-infected. We are aware of a participant who despite his family members developed Sars-Cov-2 infection during the second pandemic, he/she resulted negative on swab, never reported any clinical symptom of infection but had his/her anti-Spike titer increasing 4 times indicating a protective effect. A growing number of studies are showing that natural infection does protect against SARS-CoV-2 reinfection and/or symptomatic disease[32-35]. A recent study by Hall et al, analyzed data from 8278 individuals with known previous SARS-CoV-2 infection and positive for antibody at enrolment and 17 383 individuals who were seronegative and without past-infection with SARS-CoV-2 and found that previous SARS-CoV-2 infection provided a 84% risk reduction for reinfection and 93% risk reduction for those with symptomatic infections despite the concern of the circulation of variant of concern known as B.1.1.7[36].

### Strength and limitation

Strength of our study include follow-up that lasted for at least one year across from the first pandemic from, the use of both types of serological assays for the understanding of antibody characteristics.

We acknowledge some limitations of our study. First, our study does not have a baseline serologic testing since it was conceived in late April and it was not possible to obtain serologic testing when participants had the disease. The time from disease onset and the first clinical and serologic testing was 3 to 6 months. We believe that this could not have biased our results, however, we are unsure whether those that resulted negative at the first visit – who predominately were oligo/asymptomatic participants – could have positive result on the first visit. Second, the study lacks clinical and serological information regarding those who died during the pandemic event, hence we are unable to conclude whether quantitative serologic testing could predict survival.

### Conclusion

Beside determining the diagnosis of current and past infection, one of the major role of serological testing is to determine the immunization status with which it is possible to predict immunity from future infection. The use of SARS-CoV-2 serological tests requires understanding of how these tests perform in populations over time. Immunologic response from Sars-Cov-2 infection is characterized by both anti N and anti-S antibodies. While anti-N antibodies can be useful for diagnosis of past disease they do not persist overtime and may have any role in the protection from a reinfection. Given its qualitative characteristic it might not have an important role in the assessment of seroprevalence. Anti-S antibody titers correlates well with disease severity, persist for at least one year and most probably provide protection from reinfection.

## Data Availability

Data are available upon request

## Contributors

IA and MM conceived the original idea of the study. IA, AG, EP, SP, RA, SA, AF, EM, LA, PM, MLO, MR, MB, EB, and MM: participated in the data analysis and interpretation, drafted and critically revised the final version of the manuscript. IA, AG, EP, SP, RA, LA, PM, MR, MB, EB, and MM: supervised the laboratory analysis. IA, AG, EP, SP, RA, LA, PM, MR, MB, EB, and MM: participated in the data collection. MM is the guarantor.

## Funding

This study is supported by the Azienda USL Umbria 2.

## Competing interests

None declared.

## Collaborators

Alessandra De Masi, Manuela Costantini, Anna Chiara Lombardo, Anna Rita Vecchiarelli, Miria Flaviani, Carla Merigiola

---

STROBE Statement—Checklist of items that should be included in reports of ***cross-sectional studies***

| Checklist of items that should be included in reports of cross-sectional studies |  |  | Page No |
| --- | --- | --- | --- |
|  | Item No | Recommendation |  |
| Title and abstract | 1 | (a) Indicate the study's design with a commonly used term in the title or the abstract | 1 |
|  |  | (b) Provide in the abstract an informative and balanced summary of what was done and what was found | 1, 2 |
| Introduction |  |  |  |
| Background/rationale | 2 | Explain the scientific background and rationale for the investigation being reported | 3, 4 |
| Objectives | 3 | State specific objectives, including any prespecified hypotheses | 4, 5 |
| Methods |  |  |  |
| Study design | 4 | Present key elements of study design early in the paper | 6 |
| Setting | 5 | Describe the setting, locations, and relevant dates, including periods of recruitment, exposure, follow-up, and data collection | 6 |
| Participants | 6 | (a) Give the eligibility criteria, and the sources and methods of selection of participants | 6 |
| Variables | 7 | Clearly define all outcomes, exposures, predictors, potential confounders, and effect modifiers. Give diagnostic criteria, if applicable | 6 |
| Data sources/<br>measurement | 8* | For each variable of interest, give sources of data and details of methods of assessment (measurement). Describe comparability of assessment methods if there is more than one group | 6, 7 |
| Bias | 9 | Describe any efforts to address potential sources of bias | 7 |
| Study size | 10 | Explain how the study size was arrived at | NA all available participants were included |
| Quantitative variables | 11 | Explain how quantitative variables were handled in the analyses. If applicable, describe which groupings were chosen and why | 7 |
| Statistical methods | 12 | (a) Describe all statistical methods, including those used to control for confounding | 7 |
|  |  | (b) Describe any methods used to examine subgroups and interactions | 7 |
|  |  | (c) Explain how missing data were addressed | 7 |
|  |  | (d) If applicable, describe analytical methods taking account of sampling strategy | 7 |
|  |  | (e) Describe any sensitivity analyses | 7 |
| Results |  |  |  |
| Participants | 13* | (a) Report numbers of individuals at each stage of study—eg numbers potentially eligible, examined for eligibility, confirmed eligible, included in the study, completing follow-up, and analysed | 8 |
|  |  | (b) Give reasons for non-participation at each stage | 8 |
|  |  | (c) Consider use of a flow diagram | 8 |
| Descriptive data | 14* | (a) Give characteristics of study participants (eg demographic, clinical, social) and information on exposures and potential confounders | 8, 9 |
|  |  | (b) Indicate number of participants with missing data for each variable of interest | 8, 9 |
| Outcome data | 15* | Report numbers of outcome events or summary measures | 8, 9 |
| Main results | 16 | (a) Give unadjusted estimates and, if applicable, confounder-adjusted estimates and their precision (eg, 95% confidence interval). Make clear which confounders were adjusted for and why they were included |  |
|  |  | (b) Report category boundaries when continuous variables were categorized | na |
|  |  | (c) If relevant, consider translating estimates of relative risk into absolute risk for a meaningful time period | na |
| Other analyses | 17 | Report other analyses done—eg analyses of subgroups and interactions, and sensitivity analyses | 10-11 |
| **Discussion** |  |  |  |
| Key results | 18 | Summarise key results with reference to study objectives | 12 |
| Limitations | 19 | Discuss limitations of the study, taking into account sources of potential bias or imprecision. Discuss both direction and magnitude of any potential bias | 14-15 |
| Interpretation | 20 | Give a cautious overall interpretation of results considering objectives, limitations, multiplicity of analyses, results from similar studies, and other relevant evidence | 12-15 |
| Generalisability | 21 | Discuss the generalisability (external validity) of the study results | 15 |
| **Other information** |  |  |  |
| Funding | 22 | Give the source of funding and the role of the funders for the present study and, if applicable, for the original study on which the present article is based | 15 |

\*Give information separately for exposed and unexposed groups.
**Note:** An Explanation and Elaboration article discusses each checklist item and gives methodological background and published examples of transparent reporting. The STROBE checklist is best used in conjunction with this article (freely available on the Web sites of PLoS Medicine at http://www.plosmedicine.org/, Annals of Internal Medicine at http://www.annals.org/, and Epidemiology at http://www.epidem.com/). Information on the STROBE Initiative is available at www.strobe-statement.org.

## Notes

### Competing Interest Statement

The authors have declared no competing interest.

### Author Declarations

Comitato Etico Regionale Umbria, CER 3695/20

